# Procalcitonin Adds Limited Incremental Value to a Simple Bedside Score for Predicting Complicated Appendicitis: A Temporal Validation Study

**DOI:** 10.64898/2026.05.14.26353219

**Authors:** BoLin He, ShuBang Cheng, Min Liu, MinHui Li

**Affiliations:** Department of Gastrointestinal Surgery, The People’s Hospital of Longhua Shenzhen, Shenzhen 518109, China; Stomatology Department, The People’s Hospital of Longhua Shenzhen, Shenzhen 518109, China

**Keywords:** Acute appendicitis, Complicated appendicitis, Procalcitonin, Bedside score, Risk stratification, Temporal validation

## Abstract

**Background:** Complicated appendicitis (CA) increases morbidity and resource use.[1,2] In the emergency setting, risk stratification must rely on rapidly available data. Procalcitonin (PCT) is frequently obtained, but its incremental value beyond basic preoperative indicators remains uncertain.[5] We aimed to quantify PCT’s incremental predictive value and develop a practical bedside score with temporal validation.

**Methods:** We conducted a retrospective cohort study of consecutive laparoscopic appendectomy patients (January 2023–December 2024). CA was defined by postoperative pathology (gangrene/necrosis, perforation, or peri-appendiceal inflammation/abscess; worst-category rule). We compared a base logistic model (age, WBC, neutrophil percentage, fever, symptom-to-surgery interval, shock index) with an extended model adding log-transformed PCT. Discrimination (AUC) and calibration were assessed. Temporal validation used 2023 for development and 2024 for testing. We also created a simple bedside score using pre-specified cutoffs and evaluated CA risk across score strata in 2024.

**Results:** In the overall complete-case cohort (n=1,792), 397 patients (22.2%) had CA. Adding PCT modestly improved discrimination in the full cohort (AUC 0.673 to 0.685). For temporal validation, 2023 included 870 patients (CA 26.9%) and 2024 included 921 patients (CA 17.7%); one otherwise eligible patient lacked a usable admission year. In the 2024 test set, discrimination was 0.662 (base) vs 0.673 (base+PCT) with a non-significant AUC difference (DeLong p=0.116); calibration slopes were near 1.0. A 7-item bedside score stratified 2024 CA risk: 9.1% (score 0– 1), 14.7% (2–3), and 34.2% (⩾ 4). Using ⩾ 4 points identified a higher-risk subgroup (PPV 34.2%, NPV 87.5%, sensitivity 46.0%, specificity 81.0%).

**Conclusions:** PCT adds modest predictive information beyond simple preoperative indicators in the full cohort, but temporal validation suggests that this incremental gain is smaller and not statistically significant in later patients. A pragmatic bedside score can support CA risk stratification and prioritization in emergency care, whereas the role of routine PCT testing may be best reserved for selected situations in which uncertainty remains after initial assessment.

## Introduction

Acute appendicitis is among the most common emergency surgical diagnoses.[1,2] Complicated appendicitis (CA) is associated with higher morbidity and resource use, and early identification can support ED triage and operative prioritization.[1,2]

Even when CT is available, early bedside risk stratification remains clinically relevant for prioritization and perioperative planning in high-throughput emergency care pathways.

Current bedside tools were largely developed for diagnosing appendicitis rather than specifically stratifying CA risk, and performance for severity prediction may be insufficient for time-sensitive emergency workflows.[3,4] Procalcitonin (PCT) is frequently ordered but its incremental value beyond simple preoperative indicators remains uncertain.[5] Whether PCT adds clinically meaningful incremental value over routinely available variables in a temporally validated setting remains unclear.

We hypothesized that adding PCT would yield, at most, modest incremental discrimination beyond routine preoperative variables, and that any incremental gain would attenuate in temporal validation. This question is clinically important because biomarkers that add little predictive value may still increase cost and turnaround time in emergency workflows, while genuinely informative markers could help prioritize imaging, operative access, and perioperative planning in time-sensitive pathways. We therefore evaluated incremental value in the full cohort, performed year-based temporal validation, and assessed pragmatic bedside scores with and without PCT.

## Methods

### Study design and population

We conducted a single-center retrospective cohort study of consecutive patients undergoing laparoscopic appendectomy between January 2023 and December 2024. Data were extracted from the institutional electronic medical record and operative/anesthesia charts using a standardized template. Missing data were infrequent (3/1,794 [0.17%] had missing symptom-to-surgery interval) and were handled using complete-case analysis for prespecified predictors to maintain a transparent modeling strategy.

### Patient identification and data sources

Eligible patients were identified using electronic medical record and operative scheduling records for laparoscopic appendectomy during the study period, with confirmation of postoperative pathology reports and prespecified preoperative predictors. Data elements were abstracted from the electronic medical record and perioperative documentation using a standardized template to promote consistency and transparency. When values were missing, implausible, or discordant across sources, the originating clinical documentation was re-checked to reconcile entries. Because this was a retrospective study, residual information bias from documentation variability cannot be fully excluded.

### Outcome definition

CA was defined by postoperative pathology using a prespecified worst-category rule (gangrene/necrosis, perforation, or peri-appendiceal inflammation/abscess).

### Prediction models and temporal validation

We compared a base logistic model (age, WBC, neutrophil percentage, fever, symptom-to-surgery interval, shock index) with an extended model adding log-transformed PCT (log[PCT+0.001]); the constant was used to handle zero values and stabilize transformation in a right-skewed biomarker distribution. Temporal validation used a year-based split with 2023 for development and 2024 for testing (standardization parameters derived from 2023).

### Bedside scores

We evaluated a pragmatic 7-item bedside score (1 point each): age ⩾40 years; WBC ⩾14× 10^9/L; neutrophils ⩾85%; PCT ⩾0.10 ng/mL; fever; symptom duration ⩾24 h; shock index ⩾ 0.80. Cutoffs were prespecified for bedside usability and alignment with commonly used clinical thresholds rather than data-driven optimization. The age ⩾ 40 years threshold was selected as a simple, clinically interpretable boundary because older age is commonly associated with higher risk of complicated disease and adverse outcomes in emergency surgical presentations. To address whether PCT is necessary in a rapid triage tool, we additionally evaluated a 6-item PCT-free version of the score using the same cutoffs (i.e., the 7-item score minus the PCT item).

### Statistical analysis

Continuous variables are summarized as median [IQR] and compared using the Mann–Whitney U test; categorical variables as n (%) and compared using χ^2^ tests. We assessed discrimination (AUC; DeLong test),[8] calibration, and clinical utility (decision curve analysis).[9] Reporting follows STROBE and TRIPOD principles.[12,13]

## Results

Baseline characteristics are shown in Table 1. In the overall complete-case cohort (n=1,792), CA prevalence was 22.2%. Temporal split analyses were conducted in the subset with a usable admission year (n=1,791); counts are shown in Table 4, with a lower CA prevalence in 2024 than 2023 (17.7% vs 26.9%).

**Table 1.** Baseline characteristics in UA vs CA (complete-case cohort; n=1,792)

| Variable | UA | CA | P |
| --- | --- | --- | --- |
| Age, years | 32.0 [25.0, 40.0] | 32.0 [25.0, 41.0] | 0.957 |
| WBC, $\times 10^9/L$ | 13.65 [10.62, 16.34] | 14.92 [12.18, 17.80] | <0.001 |
| Neutrophil percentage, % | 83.5 [76.4, 88.5] | 86.0 [81.8, 90.0] | <0.001 |
| Procalcitonin, ng/mL | 0.05 [0.05, 0.07] | 0.06 [0.05, 0.43] | <0.001 |
| Preoperative fever | 340 (24.4%) | 164 (41.3%) | <0.001 |
| Symptom-to-surgery interval, h | 21.0 [10.0, 42.0] | 24.0 [15.0, 48.0] | <0.001 |
| Shock index | 0.73 [0.63, 0.84] | 0.78 [0.67, 0.93] | <0.001 |

Full-cohort regression and performance are summarized in Tables 2–3 and Figures 1–3. In temporal validation (2024 test set), discrimination was 0.662 for the base model and 0.673 for the base+PCT model (DeLong p=0.116), with similar calibration (Table 5; Figure 4).

**Table 2.** Multivariable logistic regression (Model 1 vs Model 2)

| Variable | Base model, OR<br>(95% CI) | P value | Base + PCT model,<br>OR (95% CI) | P value |
| --- | --- | --- | --- | --- |
| **Demographic** |  |  |  |  |
| Age (per SD) | 1.16 (1.02-1.31) | 0.020 | 1.13 (0.99-1.28) | 0.061 |
| **Laboratory findings** |  |  |  |  |
| White blood cell count (per SD) | 1.18 (1.04-1.35) | 0.013 | 1.19 (1.05-1.36) | 0.008 |
| Neutrophil percentage (per SD) | 1.40 (1.18-1.66) | <0.001 | 1.32 (1.11-1.57) | 0.002 |
| Procalcitonin, log-transformed | - | - | 1.18 (1.05-1.32) | 0.004 |
| (per SD) |  |  |  |  |
| **Clinical features** |  |  |  |  |
| Preoperative fever (yes vs no) | 1.60 (1.24-2.06) | <0.001 | 1.49 (1.15-1.94) | 0.003 |
| Symptom-to-surgery interval<br>(per SD) | 1.29 (1.15-1.45) | <0.001 | 1.26 (1.12-1.41) | <0.001 |
| Shock index (per SD) | 1.25 (1.10-1.42) | <0.001 | 1.20 (1.06-1.37) | 0.006 |

**Table 3:** Main performance (AUC/DeLong, calibration, Brier; optimism-corrected AUC)

| Metric | Base model | Base + PCT model |
| --- | --- | --- |
| **Dataset characteristics** |  |  |
| Total patients, n | 1792 | 1792 |
| Complicated appendicitis, n (%) | 397 (22.1) | 397 (22.1) |
| **Discrimination performance** |  |  |
| Apparent AUC | 0.673 | 0.685 |
| Bootstrap-corrected AUC* | 0.667 | 0.678 |
| ΔAUC (95% CI)<sup>†</sup> | Reference | 0.011 (0.004 to 0.018) |
| DeLong's test *P* value | Reference | 0.001 |
| **Calibration performance** |  |  |
| Brier score | 0.161 | 0.161 |
| Calibration intercept | 0.00 | 0.00 |
| Calibration slope | 1.00 | 1.00 |
| **Model characteristics** |  |  |
| Metric | Base model | Base + PCT model |
| PCT included | No | Yes (log-transformed) |

### Temporal validation and bedside tools

**Table 4:** Temporal split counts (2023 vs 2024)

| Year | Data set | Total patients, n | Patients with complications, n (%) |
| --- | --- | --- | --- |
| 2023 | Training set | 870 | 234 (26.9) |
| 2024 | Temporal validation set | 921 | 163 (17.7) |
| **Total** | **Overall cohort** | **1791** | **397 (22.2)** |

**Table 5:** Temporal validation performance (2024 test)

| Metric | Base model | Base + PCT model |
| --- | --- | --- |
| **Validation set characteristics** |  |  |
| Year | 2024 | 2024 |
| Total patients, n | 921 | 921 |
| Complicated appendicitis, n (%) | 163 (17.7) | 163 (17.7) |
| **Discrimination performance** |  |  |
| AUC (95% CI)* | 0.662 | 0.673 |
| DeLong's test P value† | Reference | 0.116 |
| **Calibration performance** |  |  |
| Brier score | 0.144 | 0.144 |
| Calibration intercept | -0.38 | -0.44 |
| Calibration slope | 1.13 | 1.07 |
| **Calibration assessment** |  |  |
| Intercept interpretation | Slight under-estimation | Slight under-estimation |
| Slope interpretation | Slight overfitting | Minimal overfitting |

**Table S1:** Incremental metrics (NRI/IDI)

| Metric | Value (95% CI) | P value |
| --- | --- | --- |
| **Dataset characteristics** |  |  |
| Total patients, n | 1792 | – |
| Complicated appendicitis, n (%) | 397 (22.1) | – |
| **Reclassification improvement** |  |  |
| Net Reclassification Improvement (NRI) | 0.048 (–0.061 to 0.156) | 0.388 |
| **Discrimination improvement** |  |  |
| Integrated Discrimination Improvement (IDI) | 0.005 (0.001 to 0.009) | 0.014 |
| **Methodological details** |  |  |
| Bootstrap resamples, n | 200 | – |
| PCT skewness* | 7.44 | – |
| PCT transformation | Log-transformed | – |

**Figure 1:**
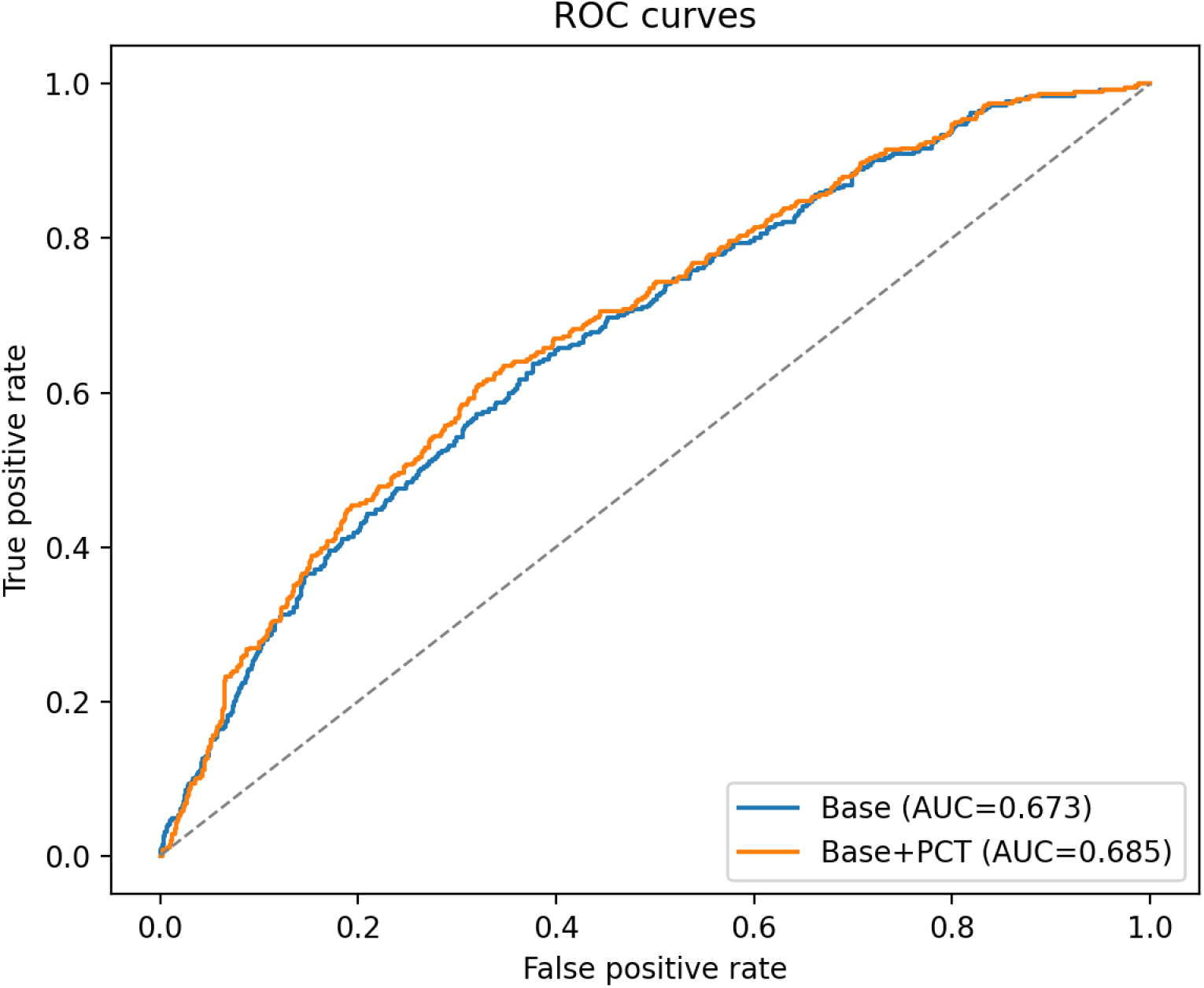
ROC (full cohort)

**Figure 2:**
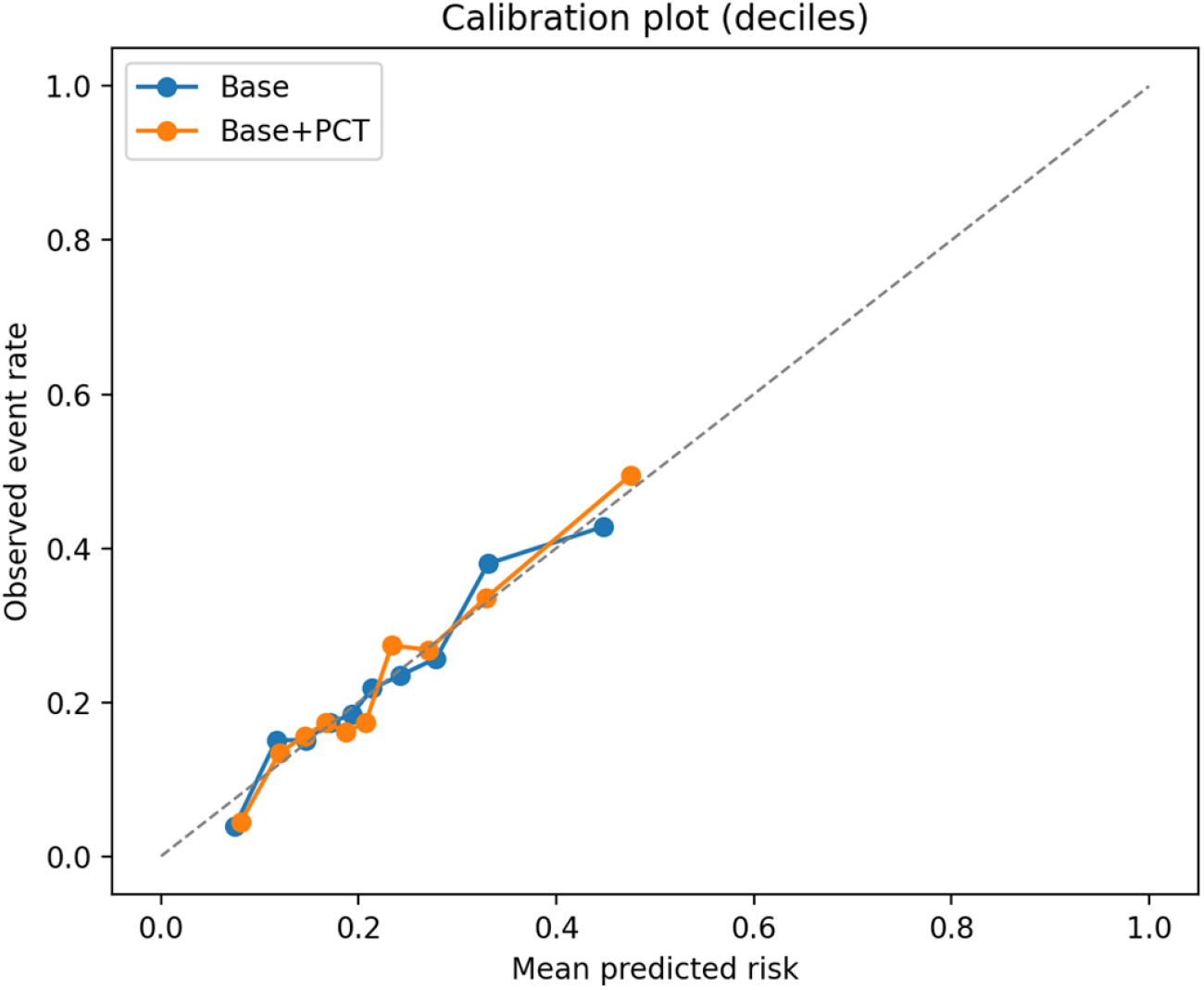
Calibration (full cohort)

**Figure 3:**
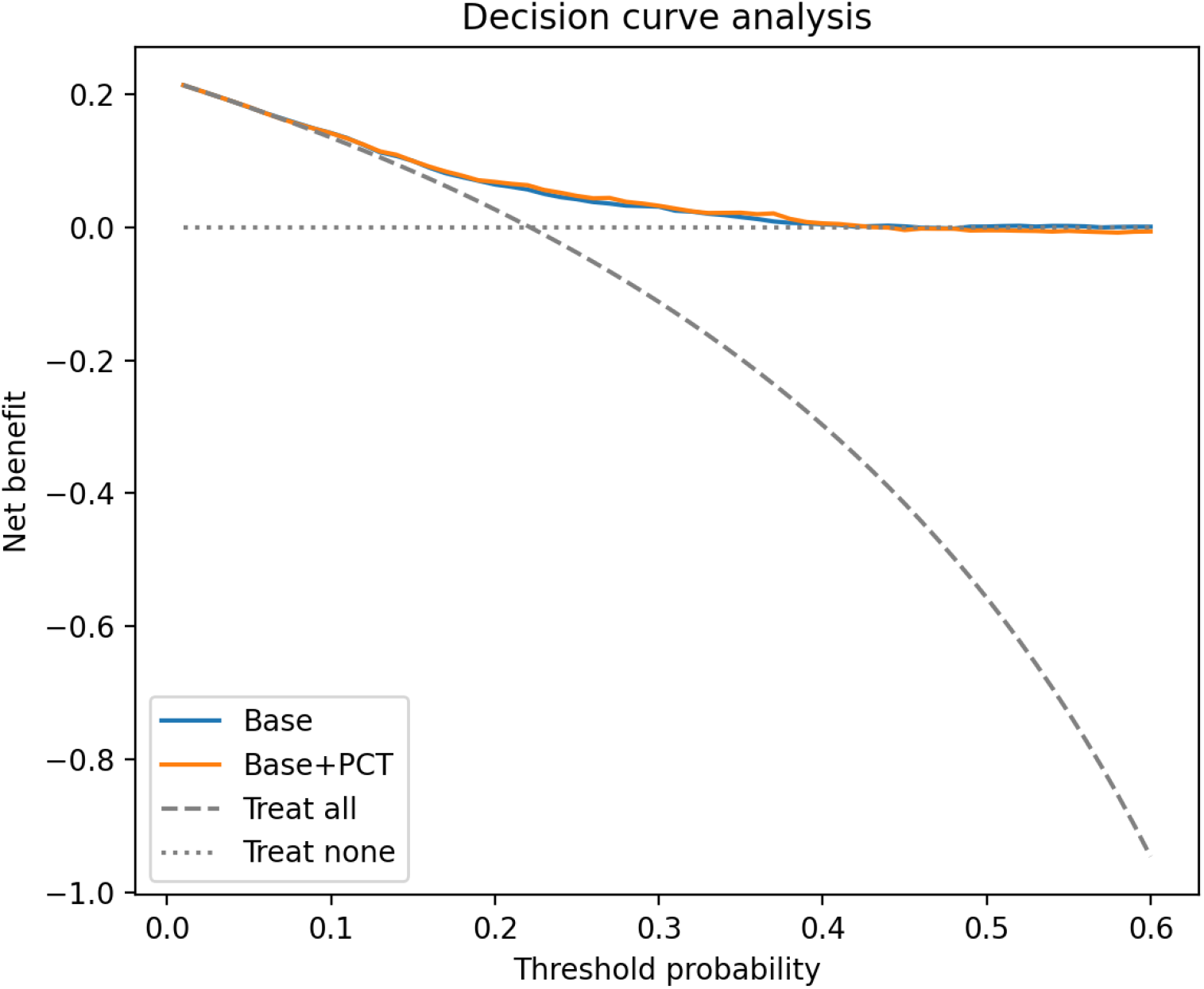
DCA (full cohort)

**Figure 4:**
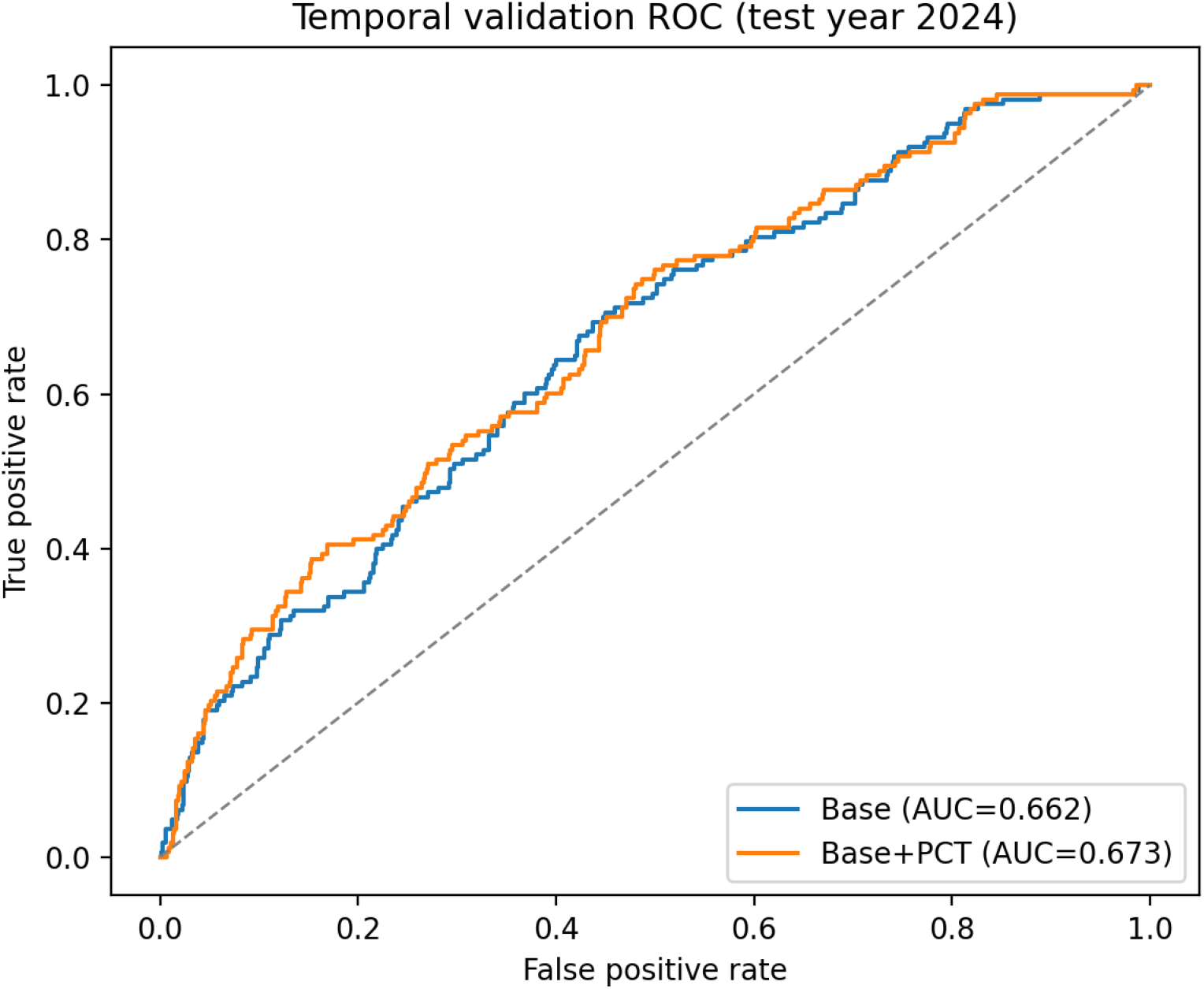
Temporal validation ROC (2024 test)

In the 2024 test set, the 7-item bedside score stratified CA risk (Table 7). Operating characteristics across thresholds for the 7-item score are shown in Table 8. To reconcile the limited incremental value of PCT with bedside implementation constraints, we also evaluated a 6-item PCT-free score, which showed similar risk stratification (Table 7) and comparable operating characteristics in the 2024 test set (Table 8). Model-based risk strata are provided as Supplementary Table S1.

**Table 6:** Bedside 7-item score definition.

| Item | Points |
| --- | --- |
| Age $\geq 40$ years | 1 |
| WBC $\geq 14 \times 10^9/L$ | 1 |
| Neutrophil percentage $\geq 85\%$ | 1 |
| PCT $\geq 0.10$ ng/mL | 1 |
| Preoperative fever ($\geq 38.0^\circ C$) | 1 |
| Symptom-to-surgery interval $\geq 24$ h | 1 |
| Shock index $\geq 0.80$ | 1 |

**Table 7:** Bedside score strata CA rates (2024)

| Score group | Total patients, n | Patients with complications, n | Complication rate, % |
| --- | --- | --- | --- |
| 0–1 (Low risk) | 274 | 25 | 9.1 |
| 2–3 (Intermediate risk) | 428 | 63 | 14.7 |
| $\geq 4$ (High risk) | 219 | 75 | 34.2 |

**Table 8:**
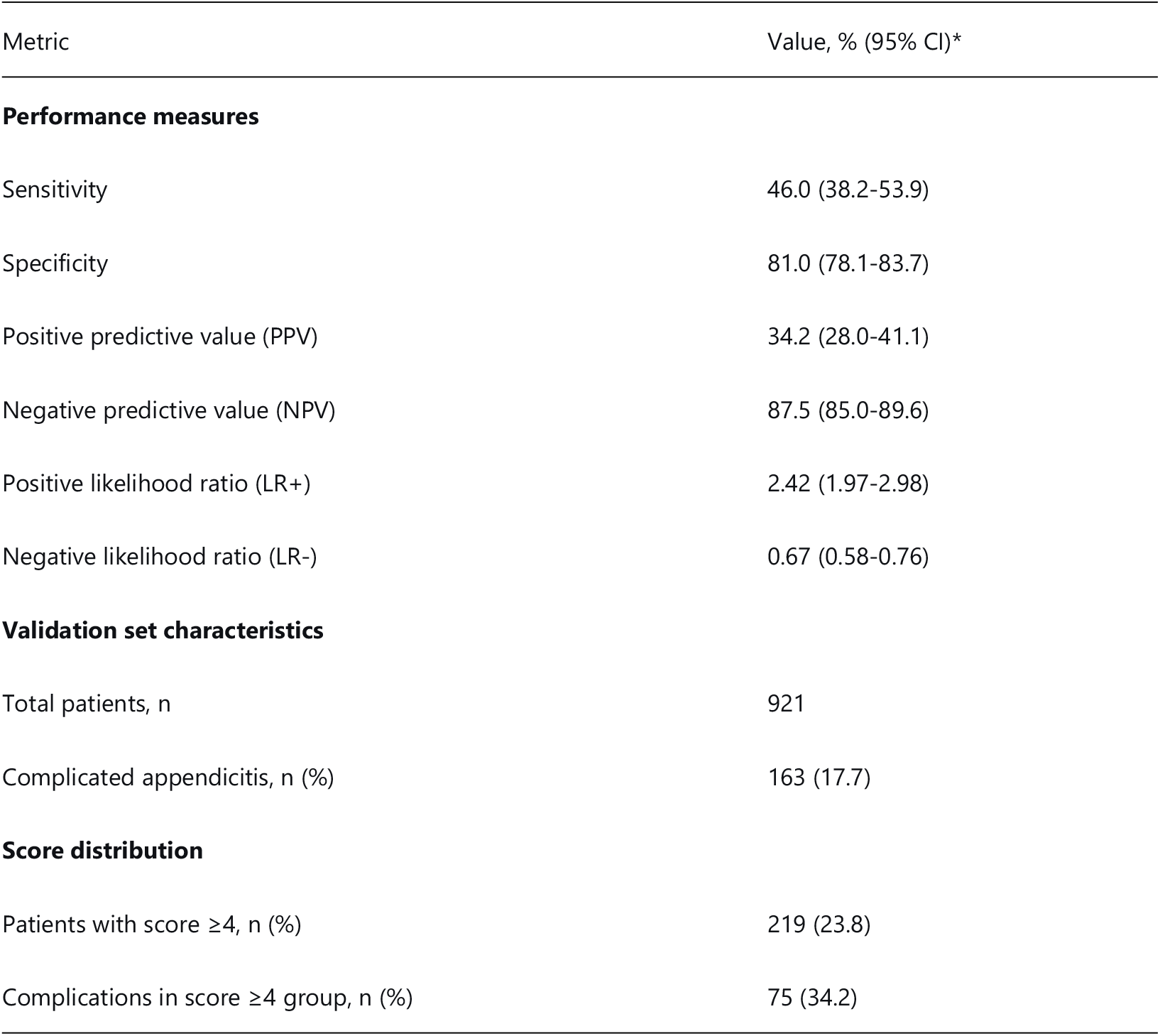
Bedside score operating characteristics (⩾4 points)

The full-cohort AUC improvement (0.673 to 0.685; Table 3; absolute Δ AUC 0.012) was statistically significant but modest, supporting cautious interpretation of clinical impact. In temporal validation, the non-significant AUC difference (DeLong p=0.116; Table 5) suggests that any incremental benefit may be limited in out-of-sample use, reinforcing that statistically detectable gains do not necessarily translate into meaningful changes in clinical triage or management decisions when workflow and cost constraints are considered.

## Discussion

### Principal findings

In this single-center cohort of appendectomy patients with pathology-defined outcomes, adding preoperative PCT to a simple base model produced a modest but statistically significant improvement in discrimination in the full cohort. In the temporally separated 2024 test set, however, the incremental gain was smaller and not statistically significant, despite similar overall calibration. These findings suggest that while PCT carries independent information related to CA risk, its added predictive contribution beyond routinely available indicators may attenuate when evaluated out of sample.

### Interpretation and the importance of temporal validation

The contrast between full-cohort performance and the time-split test set highlights a common issue in incremental biomarker research: statistical significance in model development does not necessarily translate into clinically meaningful improvement in later patients. Differences in case-mix, baseline CA prevalence, and perioperative pathways across time may further reduce incremental performance. For emergency surgery workflows, this supports the importance of temporal or external validation when assessing the practical value of candidate biomarkers and interpreting modest AUC gains cautiously.

### Clinical implications

From a practical perspective, PCT may be most useful as an adjunct in situations of residual uncertainty rather than as a routine determinant of management. Given the limited incremental gain observed in temporal validation, PCT should complement, not replace, clinical assessment and imaging when indicated. In our 2024 test set, the bedside score provided clinically relevant risk stratification, with CA rates increasing from 9.1% in the low-risk group to 34.2% in the high-risk group. At a threshold of ⩾ 4 points, the score showed moderate rule-in utility but limited sensitivity, supporting its role as a prioritization tool rather than a definitive diagnostic rule.

In busy emergency settings, a simplified PCT-free strategy may be operationally attractive because it could support earlier preliminary risk stratification without relying on additional biomarker testing. One pragmatic pathway is to use the bedside score as an early prioritization layer, for example by prioritizing higher-risk patients for imaging and operating-room access, while recognizing that delays to appendectomy and other workflow factors may influence outcomes and resource use.[6,7] In this framework, PCT may be reserved for selected patients with persistent diagnostic uncertainty, particularly when imaging is delayed or unavailable. This selective-use strategy, however, requires prospective evaluation.

### Comparison with prior work

Prior evidence supports PCT as a marker of more severe inflammation in appendicitis, but most studies have focused on diagnostic accuracy rather than incremental prediction beyond basic clinical and laboratory features.[5] Our findings align with this distinction: PCT added statistically detectable but modest discrimination in the full cohort, with attenuation in temporal validation. This pattern reinforces the need to prioritize temporal or external validation when proposing biomarkers for real-world emergency decision support.

Compared with diagnostic-oriented tools such as Alvarado and AIR,[3,4] our bedside tool is explicitly presented as a CA risk-stratification aid rather than a diagnostic replacement. Direct comparison with established scores and imaging pathways remains an important next step.

### Strengths and limitations

Strengths of this study include the large consecutive surgical cohort, pathology-based outcome definition using a prespecified worst-category rule, prespecified preoperative predictors, and a transparent temporal validation design. Limitations include the retrospective single-center setting, possible residual confounding, and potential misclassification arising from pathology text abstraction. The time-split approach does not substitute for true external validation across institutions, and complete-case analysis may introduce selection bias if missingness is not random. Future studies should determine whether simplified score-guided pathways can improve both patient-centered and operational outcomes and whether any incremental predictive gain from PCT justifies routine testing within local resource constraints.

## Data Availability

Due to patient privacy protection and institutional data governance requirements, raw clinical records containing potentially identifiable information cannot be publicly shared. De-identified patient data and complete analysis code (R version 4.2.0) are available upon reasonable request to the corresponding author, subject to a data access agreement that complies with the institutional ethics approval (Approval No. 2026-001). The authors confirm that all relevant data underlying the findings are fully available without restriction, except for privacy/ethical restrictions. Requests should be directed to the corresponding author at ? and will be responded to within 30 days. Alternative contact for data access: Institutional Ethics Committee, [The People's Hospital of Longhua Shenzhen], [NO. 38 Jinglong Construction Road].

## Ethics / Declarations

Ethics approval: This study was conducted in accordance with the Declaration of Helsinki and was approved by the institutional ethics committee (Approval No. 2026-001). Informed consent waived due to retrospective de-identified data use.

Conflicts of interest: none. Funding: none. Data availability: De-identified analysis data sufficient to reproduce the reported results, together with the analysis code/scripts used for data processing and statistical analysis, will be provided as supporting information or in a public repository at the time of submission/publication. Direct sharing of raw clinical records is restricted because of patient privacy and institutional data-governance requirements.

